# Identifying communities at risk for COVID-19-related burden across 500 U.S. Cities and within New York City

**DOI:** 10.1101/2020.12.17.20248360

**Authors:** Chirag J Patel, Andrew Deonarine, Genevieve Lyons, Chirag M Lakhani, Arjun K Manrai

## Abstract

**Background:** While it is well-known that older individuals with certain comorbidities are at highest risk for complications related to COVID-19 including hospitalization and death, we lack tools to identify communities at highest risk with fine-grained spatial and temporal resolution. Information collected at a county level obscures local risk and complex interactions between clinical comorbidities, the built environment, population factors, and other social determinants of health.

**Methods:** We develop a robust COVID-19 Community Risk Score (C-19 Risk Score) that summarizes the complex disease co-occurrences for individual census tracts with unsupervised learning, selected on their basis for association with risk for COVID complications, such as death. We mapped the C-19 Risk Score onto neighborhoods in New York City and associated the score with C-19 related death. We further predict the C-19 Risk Score using satellite imagery data to map the built environment in C-19 Risk.

**Results:** The C-19 Risk Score describes 85% of variation in co-occurrence of 15 diseases that are risk factors for COVID complications among 26K census tract neighborhoods (median population size of tracts: 4,091). The C-19 Risk Score is associated with a 40% greater risk for COVID-19 related death across NYC (April and September 2020) for a 1SD change in the score (Risk Ratio for 1SD change in C19 Risk Score: 1.4, p < .001). Satellite imagery coupled with social determinants of health explain nearly 90% of the variance in the C-19 Risk Score in the United States in held-out census tracts (R^2^ of 0.87).

**Conclusions:** The C-19 Risk Score localizes COVID-19 risk at the census tract level and predicts COVID-19 related morbidity and mortality.

## Introduction

COVID-19 has disrupted major world economies and overwhelmed hospital intensive care units around the world. In the United States alone, the virus has spread throughout urban and rural communities and killed over 300,000 Americans to date. Case-series and epidemiological surveillance data from the United States [1–4] and around the world [5–9] have implicated risk factors for COVID-19-related morbidity and mortality including older age, male sex, impaired lung function, and cardiometabolic-related diseases (e.g., diabetes, heart disease, stroke), and obesity. In the United States, comorbidities are known to “cluster” in geographies, such as the southeast states and counties (e.g., in chronic disease [10] and in COVID-19 [11–14]). Other factors including the built environment and air pollution have been associated with COVID-19 infection, complications [15,16] but it has been unclear how to prioritize these associations to prevent complications. Both individual- and geographical-level social determinant factors are strong risk factors for COVID-19 infection and risk [17].

However, even within these macro-scale hotspots, common chronic diseases and their risk factors for COVID-19 are geographically heterogeneous and vary per unit of geography, including within and across states, counties, and even cities. It is unclear how the heterogeneity of community-based risk or prevalence of diseases at a census-tract level (median population sizes of ∼3000-5000 individuals) is related to COVID-19 risk. Furthermore, analyses on coarser spatial resolutions will attenuate predictions and associations [18]. Here, we demonstrate how to calculate a COVID-19 Community Risk Score (“C-19 Risk Score”) at the census tract level that summarizes the complex co-morbidity and demographic patterns of small communities. We show how the C-19 Risk Score varies within and across cities, pointing to how county-level estimates may obscure identification of specific regions at high risk, important for resource allocation. Second, we develop a machine learning approach to trace factors from the “built environment” using deep learning methods to interpret satellite imagery -- common to those used in navigation -- to predict the C-19 Risk Score. This is inspired by work from Maharana and Nsosie, who developed an approach to map the built environment to obesity prevalence [19]. We also demonstrate how social determinants of health correlate with and predict the C-19 Risk Score. Further, we focus on a late-May 2020 hotspot of COVID-19 epidemic, New York City, to show how the C-19 Risk Score is associated with zipcode-level COVID-19 related deaths. Lastly, deploy the COVID-19 Risk Score with an application programming interface and a browsable dashboard (accessible at http://launchpad.xybeta.ai/).

## Methods

### Study Data

We obtained geocoded disease prevalence data at the census tract level from the U.S. Centers for Disease Control and Prevention 500 Cities Project (updated December 2019[20]) (Figure 1A). 500 Cities contains disease and health indicator prevalence for 26,968 number individual census tracts of the 500 Cities which are estimated from the Behavioral Risk Factor Surveillance System [21].

**Figure 1.**
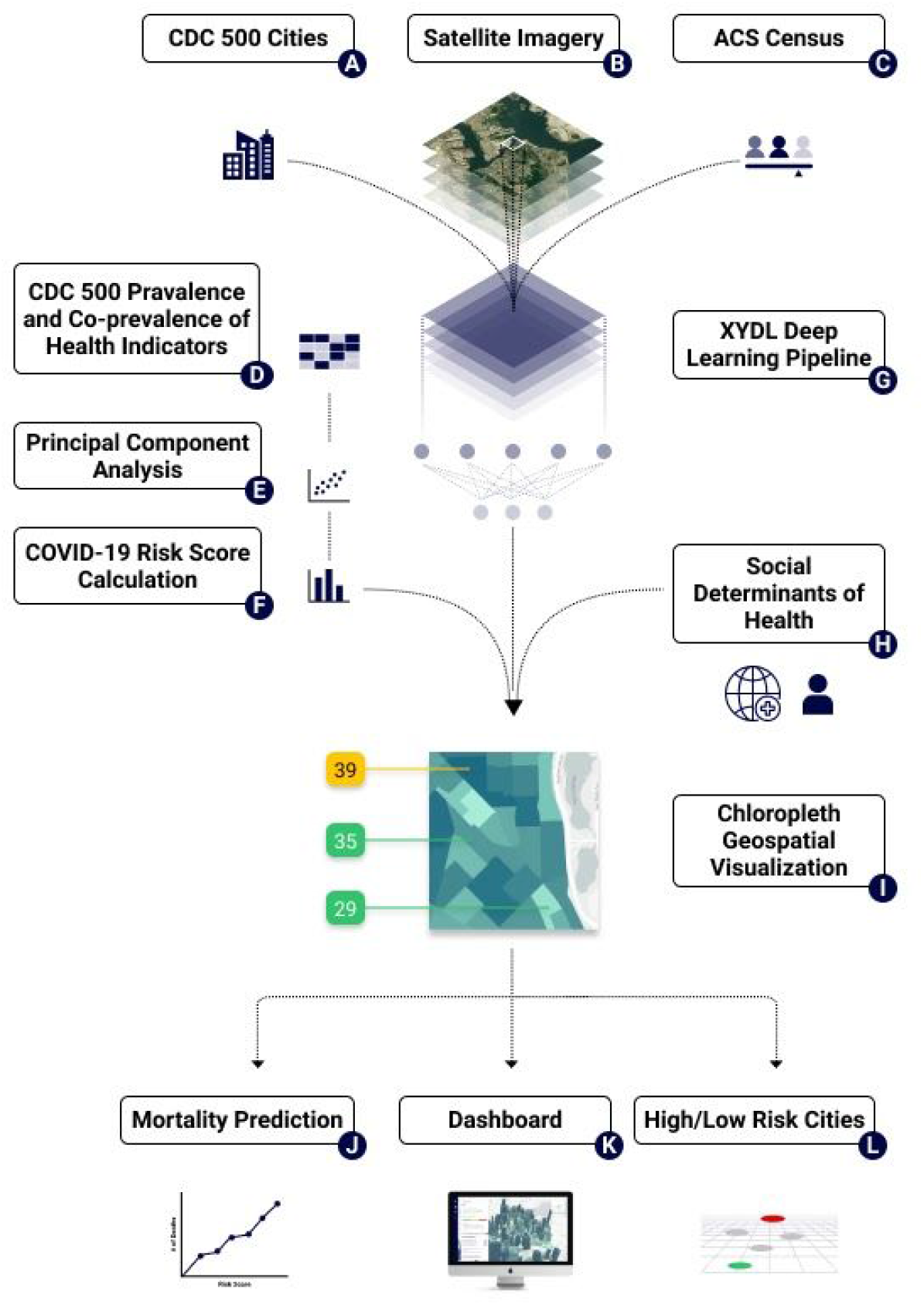
Overview of study. A.) Centers for Disease Control 500 Cities; B.) Satellite imagery of 500 cities from OpenMapTiles; C.) ACS Census summary statistics for each census tract; D.) Estimates of prevalence and co-prevalence of disease and health indicators for risk of COVID-19 Complications; E.) Use of Principal Components Analysis (PCA) to reduce dimensionality of diseases and health indicators; F.) Construction of COVID-19 Score from principal components; G.) “XYDL” deep learning pipeline that inputs satellite imagery, social determinants of health indicators from ACS Census data to predict COVID-19 Community Risk Score, H.) Social determinants of health from ACS Census data, I.) Visualization of the COVID-19 Community Risk Score, J.) Association of the COVID-19 Risk Score with mortality in NYC; K.) Creation of a dashboard; L.) Mapping highest and lowest risk cities and tracts as a function the Risk Score.

From the 500 Cities data, we chose 13 health indicators that are associated with COVID-19-related hospitalization and death based on reports from China, Italy, and the United States (e.g., [1,6,8,9]). Disease indicators include the prevalence amongst adults of diabetes, coronary heart disease, chronic kidney disease, asthma, arthritis, any cancer, chronic obstructive pulmonary disorder. We also selected behavioral risk factors including smoking and obesity. Third, we selected variables that reflected access to care, such as prevalence of individuals on blood pressure medication and high cholesterol levels.

We further obtained 5-year 2013-2017 American Community Survey (ACS) Census data [22], which contain sociodemographic prevalences and median values for Census tracts (Figure 1C). We selected the total number of individuals in the tract, proportion of males and females over the age of 65, and proportion of individuals by race. These data also included information on socioeconomic indicators including median income, the proportion of individuals living in poverty, unemployed, cohabitate with more than one individual per room, and have no health insurance.

### Defining the COVID-19 Community Risk Score (C-19 Risk Score)

We merged disease, behavior, and health access prevalence data from the 500 Cities Project for each of the 26,968 census with ACS information and calculated their Pearson pairwise correlations (Figure 1D). We considered 15 variables in total, including 13 health indicators (e.g., diseases and risk factors), and 2 demographic factors, the proportion of males and females individuals over 65 in the risk score. The disease prevalence included diabetes, coronary heart disease, any cancer, asthma, chronic obstructive pulmonary disease, arthritis; the behavioral risk factors included prevalence of obesity, smoking, high cholesterol, and high blood pressure. The clinical risk factors included the prevalence of individuals on a blood pressure medication.

To summarize the total variation of the disease prevalences in a single score, we devised 2 approaches. The first approach scaled each of the 13 health indicators plus 2 indicators of males and females over 65 (z-score transformation) prevalence by subtracting the overall average and dividing by the standard deviation of the prevalence. Then, for each census tract, the z-scores were summed and rescaled to be between 0-100. Therefore, the tracts with the highest scores have the highest “additive” prevalences for all the health indicators. We call this the additive score.

The second and primary score utilized principal components analysis (PCA), a “unsupervised” machine learning approach that transforms the data (26,968 by 15 dimensions) into a new space where each new variable is a linear combination of variables from the original dataset. PCA attempts to maximize the variance explained over the dataset in successive new variables (call them Y_1_through Y_15_) that are defined by the “components”, or linear combination of each of the original variables (e.g., health indicators) (Figure 1E). Therefore, the first variables (Y_1_, Y_2_, etc) that correspond to the first principal components in the new dataset explain the maximal amount of variation in the entire dataset. After reprojecting the census tracts on the first two principal components, call them Y_1_and Y_2_(fitting each census tract to the first two principal components), we “aligned” Y_1_and Y_2_such that the increasing prevalence of all the disease and health indicators were monotonically increasing with increase of the disease prevalences. Next, we estimated a single score for each census tract as a weighted average between Y_1_ and Y_2_, where the weights were proportional to the variance explained by the first and second principal components respectively. Finally, the score is rescaled to be between 0-100. The higher the value, the higher the total burden of disease and proportion of individuals over 65 in that census tract. We calculated risk scores for different units of administrative areas, including the 500 cities, 316 counties, and 50 states. We estimated a city-wide prevalence of each of the 15 COVID-19 risk factors and diseases and then computed the additive and PCA-based scores as above. We repeated the same procedure for counties (m=316) and states (m=50).

Next, we sought to estimate how robust the PCA-based risk score is to sampling error via simulation. To do so, we estimated the standard deviation of the prevalences as a function of the size of the population of the tract. We assumed a covariance structure between the disease prevalences to be the observed census-level correlation across the US. Next, for each census tract, we simulated 100 times the prevalence of the diseases using a multivariate normal distribution, centered around the actual prevalence and with covariance equal to the COVID-19 risk factor correlation over all 26,968 tracts. Next, for each of the 100 simulated datasets we computed the principal components and obtained a simulated distribution about the principal component. Next, we estimated the predicted new projections for plus or minus 5 standard deviations (SD) of the principal components. The “robustness” score is the range of the score across the +/- 5 SD of the principal components.

### Socioeconomic Correlates of the Community COVID-19 Risk Score

We associated each of the ACS-estimated sociodemographic indicators with the C-19 Risk Score multivariate linear and random forests regression to test the linear and non-linear contribution of the sociodemographic indicators in the COVID-19 Score (Figure 1H). We split the dataset into half “training” and “testing” to get a conservative estimate of variance explained and predictive capability of the sociodemographic variables in the COVID-19 Risk Score while not overfitting the data. Specifically, we tested the linear and non-linear association prediction between American Community Survey 5-year proportions of individuals in each census tract who were (a) below poverty, (b) unemployed, (c) non-employed, (d) have less than high school education, (e) lack health insurance, (f) have more than one person occupied per room, and (g) Hispanic, Asian, African American, or Other Ethnic group. Furthermore, we also included the median income and median home value of a census tract. Coefficients in the linear regression denoted a 1 unit change in a 1 SD change in the socioeconomic variable (e.g., a 1 SD unit change in the prevalence or a 1 SD change in the median income for a census tract). Random forests were fit using 1000 trees and a tree size of 5.

### Reconstructing the COVID-19 Community Risk Score from satellite imagery

To reconstruct the C-19 Risk Score from satellite imagery (Figure 1B), millions of satellite images (n = 4,742,919) were analyzed in an ensemble of an unsupervised deep learning algorithm and a supervised machine learning algorithm. The images are satellite raster tiles that we downloaded from the OpenMapTiles database. The images have a spatial resolution close to 20 meters per pixel allowing a maximum zoom level of 13.[23] Images were extracted in tiles from the OpenMapTiles database using the coordinate geometries of the census tracts. After extraction, images were digitally enlarged to achieve a zoom level of 18.

Many census tracts are large enough to contain multiple satellite images. The median number of images per tract is n = 94, and the number of images per census tract ranges from n = 1 image in the census tract to the largest geographical tract with n = 162,811 images (in Anchorage, AK) with an interquartile range from 43 to 182 images. The geographical coverage of the images per census tract ranges from the smallest census tract covering 0.022 km^2^ and the largest census tract covering 5,679.52 km^2^, with an interquartile range from 0.93 km^2^ to 3.89 km^2^ and a median of 1.92 km^2^ per census tract.

First, we passed images through AlexNet, a pretrained convolutional neural network, in an unsupervised deep learning approach called feature extraction.[24] (FIgure 1G) The resulting vector from this process is a “latent space feature” representation of the image comprising 4,096 features. This latent space representation is essentially an encoded (non-human readable) version of the visual patterns found in the satellite images, which, when coupled with machine learning approaches, is used to model the built environment of a given census tract.[19] For each census tract, we calculated the mean of the latent space feature representation. We performed feature extraction on a NVIDIA Tesla T4 GPU using Python 3.7.7 and the PyTorch package. Finally, the latent space feature representation was regressed against the COVID-19 Risk Score using gradient boosted decision trees.[25] We split the 80% of the data into training and the remaining 20% to testing. To train the model, we used a maximum tree depth of 5, a subsample of 80% of the features per tree, a learning rate (i.e., feature weight shrinkage for each boosting step) of 0.1, and used 3-fold cross-validation to determine the optimal number of boosted trees. Training was completed on a NVIDIA Tesla T4 GPU using Python 3.7.7 and the XGBoost package. In a separate analysis, both satellite image features and the social determinants of health features (above) were regressed against the COVID-19 Risk Score. We report R^2^ for the predictions in the test data (Figure 1GH).

### Association of the COVID-19 Community Risk Score with zipcode-level COVID-19-attributed mortality

We downloaded case and death count data on a zipcode tabulation area (ZCTA) of New York City, a hotspot of the US COVID-19 epidemic as of 5/20/20 and then again on 9/20/20 (Figure 1J). We used 2010 Census cross-over files to map Census tracts to ZCTAs. We computed the average C-19 Risk Score for the ZCTA, weighting the average by population size of the Census tract. As above, we estimated the ZCTA-level socioeconomic values and proportions. We associated the C-19 Risk Score with the death rate using a negative binomial model. We set the offset term as the logarithm of the total population size of a zipcode. The exponentiated coefficients are interpreted as the incidence rate ratio for a unit change (e.g, 1 standard deviation increase) in the variable (versus no change).

### The Community COVID-19 Risk Score Application Programming Interface and Dashboard

To distribute the data to other software applications, users, and provide interactive graphing capabilities, we created an Application Programming Interface (API) and dashboard for the C-19 Risk Score. We obtained COVID-19 case data from Johns Hopkins University[26]. The API endpoints and their outputs are described in Appendix I. We also developed a dashboard through the API in the form of geographical coordinate information for Census tracts, and data values (in GeoJSON format) to construct a choropleth in the web browser. We obtained geographical coordinates and shapefiles from the US Census (2018)[27]. Distributions of values are displayed for Census tracts being viewed at a particular location (the interactive dashboard is currently available online at https://dashboard.xyai-health.com/ (see a screenshot in the Supplementary Information). Our code and data are available as open-source under the MIT license [28]. We also have deployed an Application Programming Interface for access to the C-19 Risk Score; for more details see the Supplementary Information.

## Results

### Prevalence and heterogeneity of COVID-19 associated co-morbidities and risk factors across 500 Cities of the United States

Figure 1 describes the strategy of the study. We present summary statistics of the prevalence among the 15 candidate COVID-19 comorbidities and risk factors across all of the 29,768 census tracts across the United States using US Centers for Disease Control and Prevention (500 Cities) data from 2017 and American Community Survey data collected between 2013-2018 (Figure 1A and 1C). We chose these comorbidities and risk factors because they were (1) among the strongest risk factors for COVID-19 related hospitalization, intensive care unit utilization, and death (e.g., males and females above 65, diabetes, heart disease, and stroke), (2) were indicative of risk for cardiometabolic disease or impaired lung function (e.g., smoking, obesity, high blood pressure, high cholesterol, kidney disease, asthma, chronic pulmonary obstructive disorder), or (3) may be indicative of drug use that might impair immune systems (e.g., cancer, arthritis, blood pressure drug use).

Census tracts represent small “communities” that have a median population size of 4,091 (total range of 15-51,536). For example, of the 500 cities in our analysis, cities had a median 28 number of census tracts (IQR of 20 to 47). The cities with the largest number of tracts included New York (2,109 tracts, n=8,440,712), Los Angeles (992 tracts, n=3,961,681), Chicago (794 tracts, n=2,726,431), Houston (552 tracts, n=3,067,519), and Philadelphia (376 tracts, n=1,575,522). The cities with the smallest number of tracts included Meridian, ID (4 tracts,n=53,442), Fishers, IN (9 tracts, n=96,506), Perris, CA (9 tracts, n=87,7633), Pharr, TX (9 tracts, n=77,907), Chino Hills, CA (10 tracts, n=69,461), and Lynwood, CA (10 tracts, n=69,461).

We found that risk factor prevalence and its range in census tracts across the entire US was, as expected, large (ranging from 6% to 100% [entire tract]) (Figure 1D, Figure 2A). For example, the median proportion of females and males over that age of 65 is 14% and 11% respectively (interquartile range [IQR] of 10-18% for females and 7-15% for males). The median proportion of diabetes and heart disease was 10% (IQR of 8-13%) and 5% (IQR of 4-7%); on the other hand, the median proportion of a census tract that were on treatments for, or had risk factors for heart disease, was over a third of their respective communities. Specifically, the proportion having high blood pressure, on blood pressure lowering medication, or had high cholesterol was 30% (IQR of 25-36%), 72% (IQR of 67-76%), and 32% (IQR of 29-34%) respectively.

**Figure 2.**
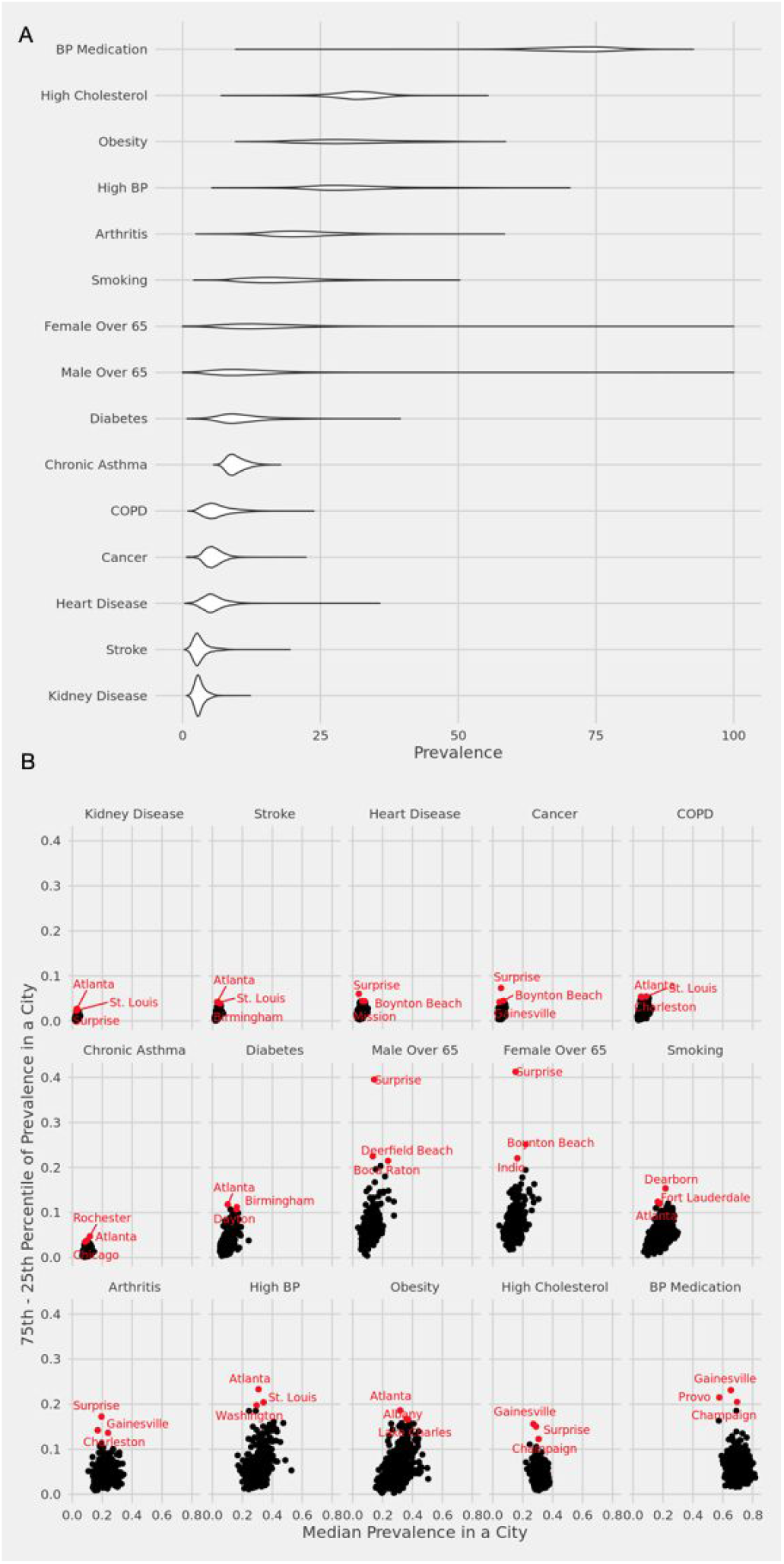
**A.) Per-census tract prevalence for health indicators (y-axis) B.) Median prevalence within a city versus the interquartile range of the prevalence of health indicators (separate panels). Top 3 cities with the largest IQR are displayed**.

The median proportion of individuals with impaired lung function, such as chronic obstructive pulmonary disorder and asthma was 10% (IQR of 6-18%) and 6% (IQR of 5-8%) respectively. Smoking, a risk factor for both impaired lung function and heart disease was 17% (IQR of 13-22%).

There was a large variation of COVID-19 comorbidities and risk factors within cities themselves (Figure 2B, Supplementary Table 1, 2) as ranked by the interquartile range (IQR, 75th percentile minus the 25th percentile) of the within city prevalence. For example, Atlanta had the greatest IQR for obesity (IQR: 22 to 40%), high blood pressure (20 to 44%), and COPD (4 to 9%), while places such as Gainesville had the highest variation in prevalence of high cholesterol (18 to 34%) and blood pressure medication (51 to 74%). Surprise, Arizona, had the highest variation in population over 65 (females:11 to 52%, males: 9 to 49%) and cancer prevalence (5 to 12%).

### Patterns in co-morbidities and risk factors across the United States

We estimated the cross-census tract correlation between the 15 COVID-19 candidate health indicators and risk factors (Figure 1D, Figure 3A). We found that there was dense correlation (median absolute value of correlation was 0.63 (IQR of 0.35 to 0.78) amongst many of the disease prevalences; specifically, higher or lower prevalence of one disease was strongly associated with higher or lower prevalence of their risk factors or complications of disease. For example, cardiometabolic diseases, such as diabetes, stroke, and heart disease displayed a strong correlation (mean pairwise Pearson correlation between these 4 disease outcomes were 0.92) between one another. Risk factors for these diseases, such as obesity, high blood pressure and high cholesterol exhibited on average a correlation of 0.62 between them, and furthermore, an average correlation of 0.78 between diseases such as diabetes, stroke, and heart disease.

**Figure 3.**
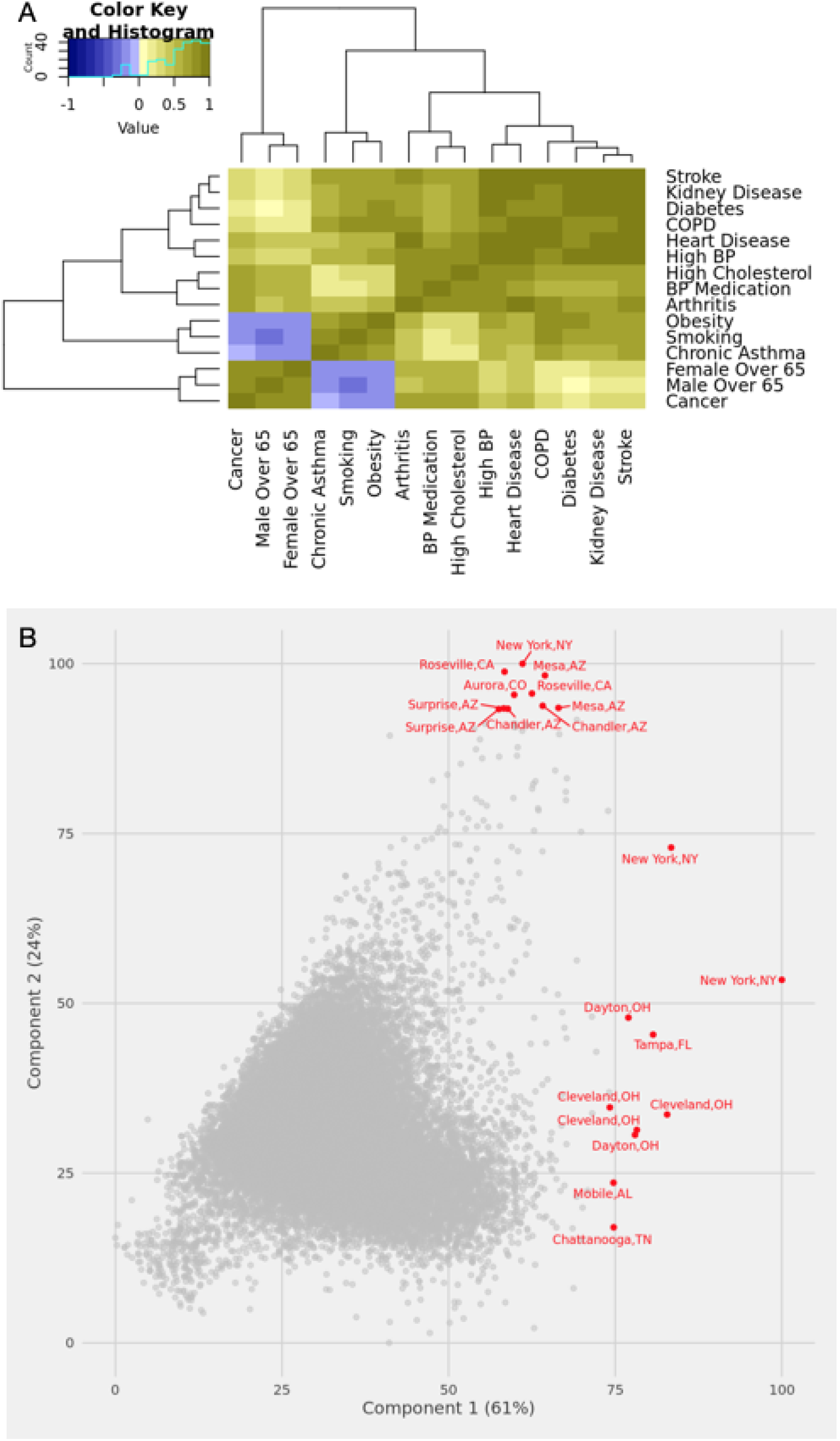
**A.) Correlation of health indicators across 27,968 census tracts. B.) Variables corresponding to the first two principal components of health indicators in the United States**

Communities that had a higher prevalence of smoking also exhibited higher prevalence of COPD and chronic asthma. The correlation between asthma and COPD was 0.69, between smoking and COPD was 0.81, and between smoking and asthma was 0.78. Obesity, an established risk factor for many diseases, was correlated with all disease indicators and risk factors (mean absolute value of pairwise correlation of 0.54). Communities that had higher prevalence of males and females older than age 65 larger prevalence of any cancer (average correlation of 0.78).

The first two principal components of the 15 COVID-19 health indicators and risk factors described 85% of the total variation (61 and 24% for component 1 and 2, respectively) of the variation over all 29,768 census tracts (Figure 1E). The first principal component had equal contribution from all 15 health diseases, except for cancer and males and females over the age 65; the second principal component was dominated by cancer and age (Figure 3B, Supplementary Table 3). The structure across health indicators and risk factors persisted when examining less specific geographical units of counties and states.

### Calculating a robust COVID-19 Community Risk Score

To estimate a C-19 Risk Score, we reprojected the 29,978 tracts onto the first two principal components, reducing the 15 health indicators to two variables (Figure 1EF). The COVID-19 score was the weighted sum of the two variables, where the weight was the total variance explained for that variable (61% and 24% respectively). We scaled the score to be between 0-100. The average score was 33.7 (SD: 8.6); the median of the score was 33.32 and the interquartile range was 28 to 38. Table 1 shows the communities with the highest variation of scores in the US.

**Table 1.** Cities with the largest variation of C19 Risk Score. Pctile: Percentile, SD: standard deviation, IQR: interquartile range

| Citiy, State | median | min | max | 25th pctile | 75th pctile | SD | IQR |
| --- | --- | --- | --- | --- | --- | --- | --- |
| Athens,GA | 32.2 | 6.3 | 42.6 | 20.7 | 35.7 | 10.0 | 15.0 |
| Atlanta,GA | 33.7 | 4.7 | 53.7 | 23.8 | 41.4 | 11.0 | 17.6 |
| Boynton Beach,FL | 41.7 | 21.6 | 81.3 | 35.5 | 52.1 | 16.2 | 16.6 |
| Champaign,IL | 29.8 | 3.4 | 45.2 | 17.9 | 34.1 | 12.8 | 16.2 |
| Gainesville,FL | 27.1 | 2.2 | 75.2 | 16.6 | 38.5 | 15.0 | 21.9 |
| Hemet,CA | 41.2 | 30.1 | 67.9 | 36.4 | 53.4 | 11.1 | 17.0 |
| Mesa,AZ | 32.2 | 7.7 | 84.6 | 29.0 | 43.2 | 14.7 | 14.2 |
| Montgomery,AL | 41.0 | 15.0 | 61.3 | 33.5 | 47.6 | 9.8 | 14.1 |
| St. Louis,MO | 36.9 | 22.0 | 57.3 | 31.7 | 46.3 | 8.6 | 14.6 |
| Surprise,AZ | 30.2 | 24.1 | 77.9 | 26.1 | 58.7 | 20.6 | 32.6 |
| Birmingham,AL | 43.9 | 18.8 | 57.9 | 36.4 | 49.3 | 9.6 | 12.9 |
| Cape Coral,FL | 43.4 | 30.3 | 63.3 | 37.3 | 49.3 | 8.2 | 12.0 |
| Clearwater,FL | 42.9 | 28.7 | 66.4 | 39.6 | 51.5 | 8.0 | 12.0 |
| Cleveland,OH | 42.4 | 18.3 | 78.3 | 38.0 | 49.6 | 9.1 | 11.6 |
| Dayton,OH | 43.1 | 6.0 | 78.1 | 38.6 | 49.8 | 11.8 | 11.2 |
| Huntsville,AL | 42.7 | 22.1 | 56.3 | 32.9 | 45.6 | 8.4 | 12.7 |
| Lake Charles,LA | 41.5 | 27.4 | 54.0 | 36.4 | 46.5 | 7.5 | 10.1 |
| Lakeland,FL | 43.9 | 18.0 | 65.8 | 38.3 | 49.2 | 10.9 | 10.9 |
| Largo,FL | 45.0 | 26.1 | 75.3 | 41.1 | 53.2 | 11.6 | 12.2 |
| Palm Coast,FL | 46.8 | 33.9 | 58.1 | 43.7 | 54.8 | 7.5 | 11.0 |
| Pompano Beach,FL | 43.6 | 27.1 | 64.3 | 37.0 | 48.5 | 9.7 | 11.5 |
| Shreveport,LA | 42.8 | 21.7 | 64.0 | 37.7 | 49.7 | 8.6 | 12.1 |
| Gary,IN | 50.8 | 42.5 | 61.8 | 47.1 | 54.6 | 5.2 | 7.6 |

Through simulations of the co-prevalences of each of the 29,978 census tracts, we found that the point estimates for the community risk scores were robust to simulated sampling error. The average error of the C-19 Risk Score across 29,768 tracts was 1.25 (SD of 0.85).

Many cities in the Southwest and Southeast demonstrated large disparities in the COVID-19 risk score (Figure 3C). For example, Surprise, Arizona, had C-19 Risk Scores IQR of 26 to 59. Atlanta, Georgia, had an IQR of 24 to 41.

### The COVID-19 Community Risk Score can be explained by social determinants of health and satellite images of the built environment

“Social determinants of health” and demographic characteristics of a community (Figure 1C, 1H) explain a 54% of the total additive variation of the C-19 Risk Score in the testing dataset. Second, we found an additional 11% of variation attributed to non-linear relationships, or a total of 65% between social determinants and the C-19 Risk Score in the testing data using random forest based regression. Third, we found that features of the built environment captured by satellite images contributed to 27% of the variation in the C-19 Risk Score. In total, combining both social determinants and satellite imagery explained 87% of the variation of the C-19 Tisk Score (Figure 1G, 1H). Below, we describe the most important features of each of the models.

All 13 sociodemographic variables contribute independently to the C-19 Risk Score (linear regression p-values less than 2×10^−16^ for 11 out of 13 variables); however, the relationships are complex (Table 2). The variables that had the largest additive contribution included the proportion of the community that was non-employed (for a 1 SD change in proportion of non-employed was associated with a 5.3 unit increase in the COVID-19 Score, *P* < .001). Similarly and independently, a 1 SD increase in the increase of individuals with less than a high school education was associated with a 2 unit increase in the score. However, for a 1 SD change in the increase of those at or below the poverty level was associated with a 3.3 unit decrease in the COVID-19 Risk Score.

**Table 2.**
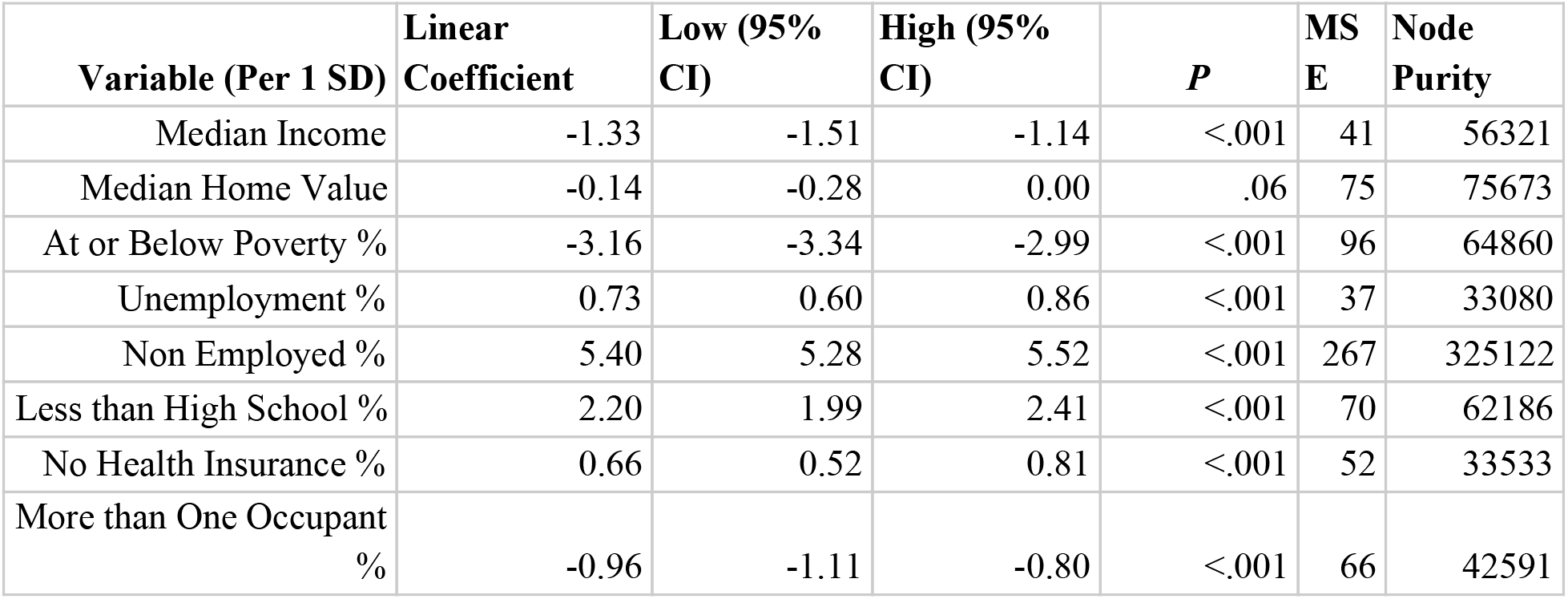
**Multivariate coefficients and confidence intervals for linear regression; mean standard error and regression impurity (residual sum of squares) for random forest for prediction of the C19 Risk Score**.

We trained a random forest regression to test for non-linear relationships (1000 trees and 4 variables at each split) in the training data. In the testing data, the social determinants of health indicators explained 65% of variation in the C-19 Risk Score in the US. The “most important” variables in the training data, ascertained through a permutation of each variable sequentially, included the proportion of the tract that was not employed (273% increase of mean-squared error [MSE] when permuted), of Asian ethnicity (93% increase of MSE), at or below poverty (91% increase of MSE), Hispanic (78% increase MSE), and less than high school (78% increase MSE). The rank order of the importance of these variables was similar to the strength of their association in the linear model (Table 2).

We hypothesized that the C-19 Risk Score is highly correlated with indicators of the built environment as captured by satellite imagery, even after accounting for social determinants of health. We deployed existing deep learning models originally trained on images from the internet and “retrained” them to predict C-19 Risk Score. In testing data, satellite imagery information plus 13 social determinants indicators explained 87% of the C-19 Risk Score variation. We deploy our findings as visualizations in a dashboard (Figure 1IJK).

### COVID-19 Community Risk Score is associated with COVID-19 death rate in New York City

Neighborhoods with higher C-19 Risk were associated with higher rates of COVID-19 related deaths in New York City (Figure 1L). We mapped the C-19 Risk Score to each zipcode tabulation area (ZCTA) in New York City in April and September 2020. Each ZCTA had information on the total number of COVID-19 tests, positive cases, and COVID-19 related deaths. We associated the C-19 Risk Score with the total death count of each zipcode, adjusting for social determinants of health variables. A 1 SD increase in the COVID-19 Risk Score was associated with a 40% increase in the incident rate ratio (Incidence Rate Ratio [IRR] of 1.40 per 1SD increase, *P* < .001) (Figure 4, Table 3) in both May and September. For zipcodes (e.g., Figure 4 annotated zipcodes) that had C-19 Risk Scores greater than 40 had an almost 2 fold increase in death rates (IRR of 1.98, 95 %CI: [1.43, 2.77], *P* < .001)

**Table 3.** **Multivariate incidence rate ratios (for 1 standard deviation change in the variable) for zipcode-level deaths in New York City in April and September 2020. IRR: incidence rate ratio. SD: Standard deviation**

| Variable (per 1 SD unit) | Apr. IRR (95% CI) | Apr. *P* | Sept. IRR (95% CI) | Sept. *P* |
| --- | --- | --- | --- | --- |
| C-19 Risk Score | 1.40 (1.27,1.55) | <.001 | 1.40 (1.27, 1.53) | <.001 |
| Median Income | 1.02 (.84,1.22) | .8 | .99 (.82, 1.18) | .9 |
| Less Than High School | .81 (.008, 1.81) | .1 | .81 (.62, 1.06) | .1 |
| College Educated | .93 (0.26, 1.92) | .5 | .93 (.76, 1.14) | .5 |
| African American | 1.14 (1.03, 2.78) | .03 | 1.16 (1.03,1.30) | .01 |
| Mexican | .9 (.87,1.08) | .6 | .97 (.87,1.07) | .5 |
| Hispanic | 1.27 (1.19,1.46) | <.001 | 1.29 (1.12,1.47) | <.001 |
| Asian | 1.12 (1.00,1.26) | .05 | 1.15 (1.02,1.28) | .02 |
| At or Below Poverty | 1.04 (.87,1.25) | .6 | .99 (.83,1.17) | .9 |
| More than 1 occupant per room | 1.12 (1.00, 1.27) | .06 | 1.10 (.98,1.23) | .1 |
| No Health Insurance | 1.02 (.91,1.16) | .7 | 1.03 (.91,1.16) | .7 |
| Unemployment | 1.01 (.91,1.13) | .8 | 1.02 (.91, 1.14) | .7 |
| COVID Case Count | 1.08 (.97,1.21) | .1 | 1.09 (.98,1.21) | .1 |

**Figure 4.**
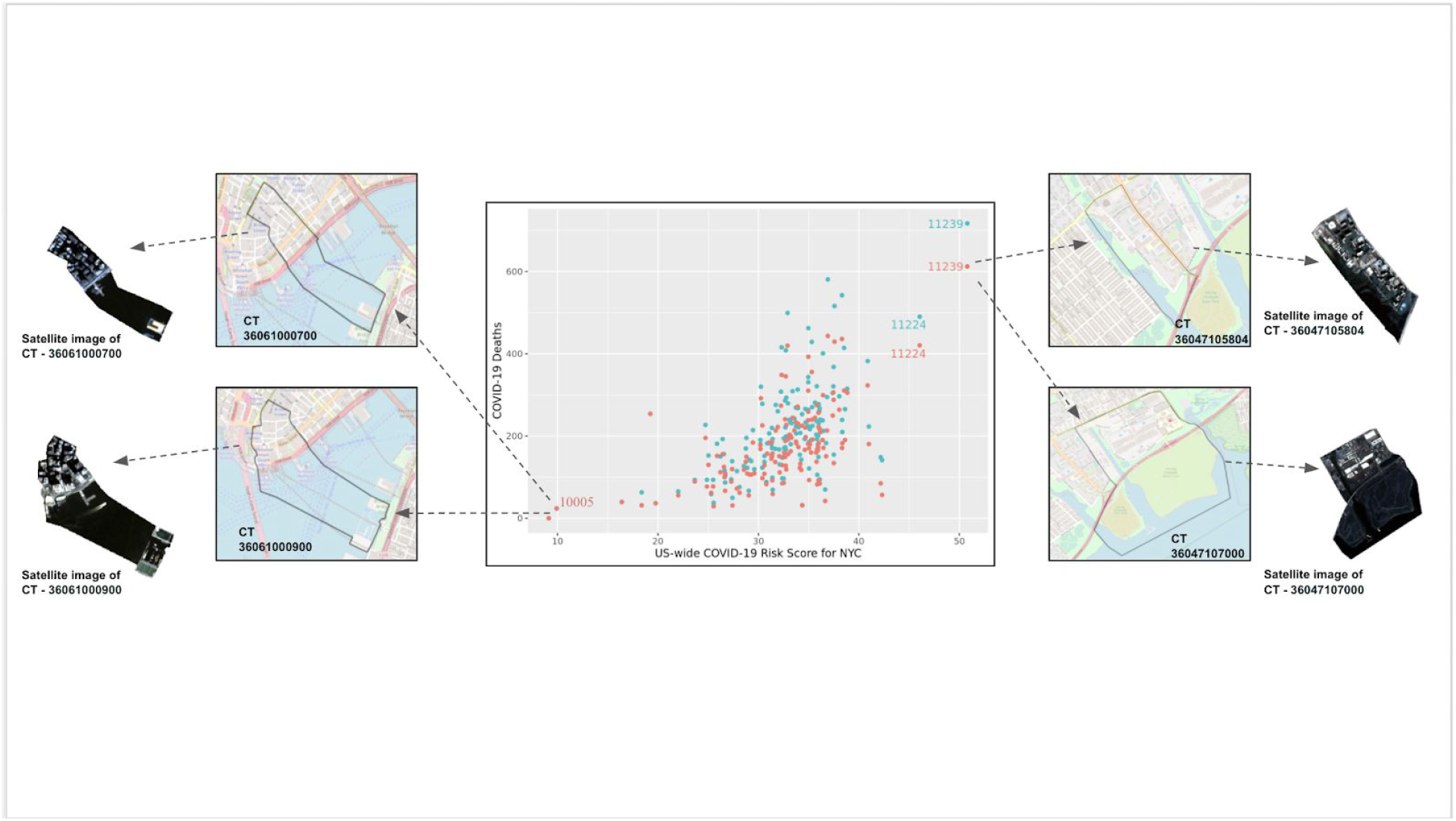
**Correlation of death rates (middle panel) and C19 Risk Score in New York City for each zipcode. The zipcodes with the highest and lowest death rates are annotated. Blue points denote data the epidemic death counts in September 2020. Red points denote epidemic death counts in April 2020**.

## Discussion

### Principal Results

In this multi-scale analysis integrating spatial disease information from gold standard disease prevalence sources such as the US Centers for Disease Control and Prevention, social determinants of health information from the US Census, and satellite imagery data, we demonstrate an approach to identify characteristics of communities at risk for COVID-19 complications. We use the tools of unsupervised learning to develop a C-19 Risk Score that provides a single interpretable number that summarize a communities’ (census tract) aggregate risk. The constituents of the C-19 Risk Score are established neighborhood level risk factors for COVID-19, such as age, obesity, diabetes, and heart disease.

We believe that the COVID-19 Risk Score can be a tool in the growing armamentarium for public health and healthcare companies’ toolbox to enable communities to prepare for the potential onslaught of cases in the coming winter months, ultimately helping to “flatten the curve” [29] to achieve precision public health. Notably, we found that the zipcode-level COVID-19 Risk Score for New York City and surrounding areas predicted risk for COVID-19 complications, such as death. Zipcodes with the highest COVID-19 scores (in the top 5%) had double the risk of COVID-19 death versus zipcodes with lowest scores. As of this writing, New York City is contemplating another lockdown due to a surge in the very same zipcodes we identified as high risk.

As a byproduct of developing a risk score for communities, we observed that there is a great disparity of chronic disease prevalence within cities and across cities in the United States. With the exception of New York City and a few other places in the United States, public health agencies mostly report COVID-19 case and death records are collected at the level of the county. However, the findings in our study implicate that smaller populations are at risk, and counties are heterogeneous.

Furthermore, we demonstrated how COVID-19 is a disease inextricably connected to social determinants. Social determinants of health are many fold, comprising both non-modifiable and modifiable factors socioeconomic status, residential location, type of occupation, access to healthcare, physical exposures (e.g. air pollution and individual factors of the exposome [30–33]), and built environment [34]. They are hierarchical in structure and distributed over both geographic space and time whose measurement can occur on both the individual-level (exposure of a person) or area level (exposure levels of a place). Satellite images provide a microscope into the area-level “built environment”, a concept that encapsulates the physical structures of how humans live, such as the city layout, resource presence, and landscape.

We uncovered immense structure in the disease prevalence information and the correlation between disease prevalence was large. ∼90% of the variation of prevalence of the 15 disease and health indicator prevalences (e.g. diabetes, obesity, cardiovascular disease, populations that take blood pressure medication, and average age, among others) can be explained by just two dimensions.

## Limitations

There are a few limitations of our study. First, we rely on disease and health indicator prevalence from the 500 largest cities in the United States, but miss out on less urban areas whose populations are at risk for COVID-19 complications. In the future, we aim to task satellite imaging technology to locations that cannot be covered by resource-limited public surveillance programs. Second, while the CDC 500 Cities data are reflective of the diversity of individuals who live in a Census tract, they are updated every two years and are dated to the latest collection (2019 data release reflects disease prevalence in 2017). Relatedly, individual-level disease nor COVID-19 status of individuals from these communities are measured. Last, satellite image data are captured at a resolution of approximately 20m per pixel. It is not clear from our study if higher resolution images (that can theoretically capture more human-visible details of the built environment) would lead to better predictions of the C-19 Risk Score.

It is clear that COVID-19 is a disease of disparity; however, we cannot make a causal claim between the instruments, such as the C-19 Risk Score, satellite imagery, and census-tract-level sociodemographic factors and eventual individual-level COVID-19 related complications.

### Comparison with Prior Work

Others have deployed similar risk scores to identify communities at risk[35]. Furthermore, we were inspired by the work of others that demonstrate how remote sensing images predict obesity prevalence [19]. However, to our knowledge, this is the first study to integrate with social determinants of health and satellite image information for prediction of C-19 Risk in neighborhoods. We found that, by combining established social determinants, information measured on earth with the built environment from space can explain most of the variation in the C-19 Risk Score. A mere 13 sociodemographic variables explain 50% of variation of the C-19 Risk Score. Combined with satellite images, we could identify ∼90% of variation in the C-19 Risk Score. Therefore, it may be possible to survey populations where COVID-19 testing and resource allocation is overlooked through the use of our predictors.

## Conclusions

While it is clear that individual-level comorbidities are associated with risk for COVID-19, here we show that communities’ clinical co-prevalence structure, satellite indicators of the built environment, and social determinants are predictive of risk. We provide all our tools to monitor these phenomena in real-time (https://dashboard.xyai-health.com/)

## Data Availability

CDC 500 Cities and US ACS Census are publicly available.
Satellite image data are available upon reasonable request.
All of our code and Risk Scores are publicly available and open source (MIT license): https://github.com/xyhealth/covid_comorbidity_score

## Supplementary Information, Figures, and Tables

**Supplementary Table 1. Distribution of C19 Risk Score diseases among 500 Cities census tracts**

**Supplementary Table 2. Distribution of C19 Risk Score diseases for each of the 500 Cities**

**Supplementary Table 3. Principal component loadings for the first two principal components of the C19 Risk constituent diseases and health indicators**

**Supplementary Figure 1. City-level Median C19 Risk Score versus difference in 75th percentile vs. 25th percentile C19 Risk Score**.

**Appendix. Application Programming Interface Endpoints and COVID-19 Community Risk Score Web-Based Dashboard**

## Acknowledgements

We thank Emmanuel Coloma and Sumeet Parekh for their help in crafting the figures. This study is funded by XY Health, Inc, a company that develops machine learning approaches for prediction of health outcomes using satellite and land sensor data.

## Abbreviations

CDC: US Center for Disease Control and Prevention
C19: COVID-19
PCA: Principal Components Analysis
IRR: Incidence Rate Ratio

