## Supplementary Information for "Identifying communities at risk for COVID-19-related burden across 500 U.S. Cities and within New York City"

Supplementary Information, Figures, and Tables

**Supplementary Table 1. Distribution of C19 Risk Score constituent diseases and health indicators among 500 Cities census tracts. Pct: percentile. SD: Standard Deviation**

**Supplementary Table 2. Distribution of C19 Risk Score constituent diseases and health indicators for each of the 500 Cities. Pct: percentile. SD: Standard Deviation.**
Located online here:
**https://docs.google.com/spreadsheets/d/1mEgyBQkqSOeb9qQ1lj9tTkQWqdkL_38RigIDFxO_bJg/edit?usp=sharing**

**Supplementary Table 3. Principal component loadings for the first two principal components of the C19 Risk Score constituent diseases and health indicators**

**Supplementary Figure 1. City-level Median C19 Risk Score versus difference in 75th percentile vs. 25th percentile C19 Risk Score.**

**Appendix. Application Programming Interface Endpoints and COVID-19 Community Risk Score Web-Based Dashboard**

**Supplementary Table 1. Distribution of C19 Risk Score health indicators among 500 Cities census tracts. Pct: percentile. SD: Standard Deviation**

| Health Indicator | Median | Low | High | 25 Pct | 75 Pct | SD |
|---|---|---|---|---|---|---|
| BP Medication | 0.718 | 0.096 | 0.927 | 0.668 | 0.759 | 0.079 |
| High Cholesterol | 0.317 | 0.07 | 0.554 | 0.288 | 0.344 | 0.049 |
| Obesity | 0.296 | 0.096 | 0.586 | 0.245 | 0.357 | 0.082 |
| High BP | 0.295 | 0.053 | 0.703 | 0.253 | 0.349 | 0.083 |
| Arthritis | 0.21 | 0.024 | 0.584 | 0.174 | 0.251 | 0.060 |
| Smoking | 0.171 | 0.020 | 0.503 | 0.134 | 0.217 | 0.061 |
| Female Over 65 | 0.137 | 0.000 | 1.000 | 0.096 | 0.187 | 0.079 |
| Male Over 65 | 0.107 | 0.000 | 1.000 | 0.074 | 0.149 | 0.070 |
| Diabetes | 0.101 | 0.008 | 0.395 | 0.079 | 0.13 | 0.043 |
| Chronic Asthma | 0.095 | 0.056 | 0.178 | 0.085 | 0.108 | 0.017 |
| COPD | 0.058 | 0.01 | 0.238 | 0.045 | 0.076 | 0.025 |
| Cancer | 0.055 | 0.007 | 0.224 | 0.045 | 0.066 | 0.018 |
| Heart Disease | 0.053 | 0.004 | 0.358 | 0.042 | 0.067 | 0.021 |
| Kidney Disease | 0.029 | 0.007 | 0.123 | 0.025 | 0.036 | 0.010 |
| Stroke | 0.029 | 0.003 | 0.195 | 0.023 | 0.038 | 0.015 |

**Supplementary Table 3. Principal component loadings for the first two principal components of the C19 Risk constituent diseases and health indicators**

| Health Indicator | Rotation 1 | Rotation 2 |
|---|---|---|
| Cancer | 0.16 | -0.43 |
| Arthritis | 0.3 | -0.13 |
| Stroke | 0.31 | 0.09 |
| Asthma | 0.2 | 0.3 |
| COPD | 0.3 | 0.12 |
| Heart Disease | 0.32 | -0.05 |
| Diabetes | 0.3 | 0.13 |
| Kidney Disease | 0.31 | 0.07 |
| Blood pressure medication | 0.26 | -0.22 |
| Smoking | 0.21 | 0.36 |
| High Blood Pressure | 0.32 | 0.02 |
| Obesity | 0.24 | 0.31 |
| High Cholesterol | 0.27 | -0.21 |
| Male > 65 | 0.12 | -0.42 |
| Female > 65 | 0.14 | -0.41 |

**Supplementary Figure 1. City-level Median C19 Risk Score versus difference in 75th percentile vs. 25th percentile C19 Risk Score.**

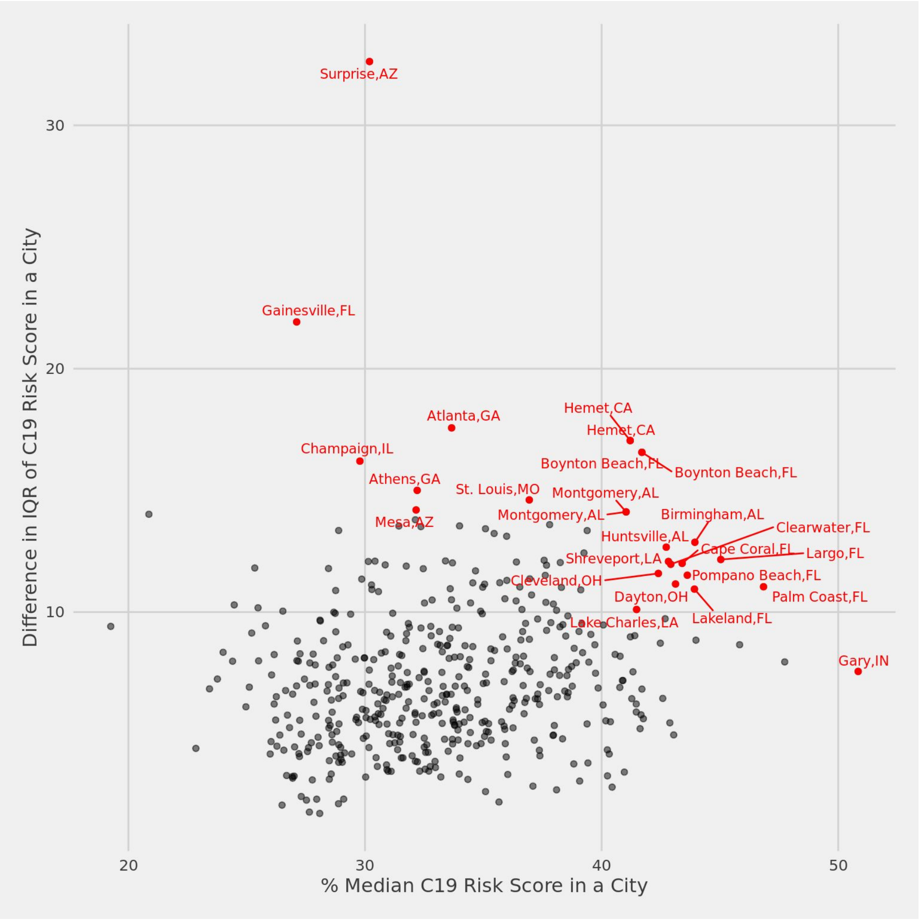

### Appendix.

### Application Programming Interface (API) Specification for the COVID-19 Risk Score

#### 1. Overview

The XY.ai COVID-19 Community Risk Score is a population-level model that ranks communities by their risk of COVID-19-related morbidity and mortality using chronic disease prevalence and demographic characteristics, summarizing information from ~134 million people. The risk score is accessed by geographical indicators, such as state, city, county, FIPS, and zip code. The COVID-19 Community Risk Score can be used to help predict community-level burden of disease, serve as a prior probability in individual-level risk models, and assist with resource allocation efforts.

The API (version 0.7) includes the XY.ai COVID-19 Community Risk Score (modelled at the state, city, county, and census tract level), underlying disease prevalence data from the CDC (census-tract estimates derived from the Behavioral Risk Factor Surveillance System (BRFSS) of the CDC 500 Cities Project, and demographic data from the U.S. Census/American Community Survey.

#### 2. Dashboard and API Specification

#### 2.1 Interactive Online COVID-19 Community Risk Score Dashboard

The interactive dashboard can now be found at the following URL:

https://dashboard.xyai-health.com

Information is displayed in interactive choropleths, as well as histograms and graphs that can be toggled on and off, allowing the user to quickly change between different datasets on the map. Additionally, summary statistics for cities can be retrieved and visualized, and COVID-19 cases are graphed as a function of the COVID-19 Community Risk score.

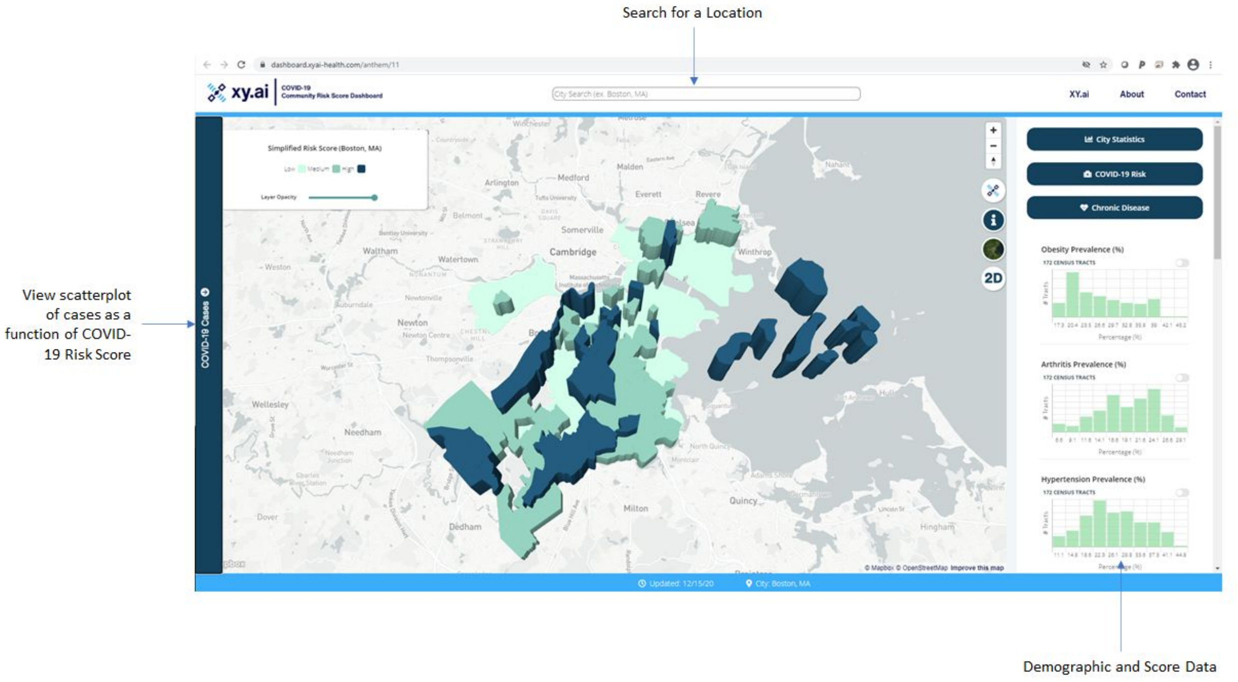


The interactive dashboard, accessed at [https://dashboard.xyai-health.com](https://dashboard.xyai-health.com).

#### 2.2 API Specification

The API is now deployed at the following URL:

https://api.xyai-health.com/{endpoint}

The API accepts JSON (POST only, GET has been disabled) parameters and returns JSON output. The current endpoints are as follows:

| **Endpoint**<br>**https://api.xyai-health.com** | **Parameters (POST)** | **Output** |
|---|---|---|
| / | None | Displays interactive dashboard |
| /status | None | Should return "OK" if the API is up |
| /state | JSON:<br>{“value”: “**stateval**”}<br><br>stateval: MA, TX…<br>or a state name (Maine, Texas, etc)<br>or state FIPS code (01, 02, 04…) | See JSON Output Object (v0.7) below |
| /city | JSON:<br>{“value”: “**cityval**”} | See JSON Output Object (v0.7) below |

| | | |
|---|---|---|
| | cityval: Boston, Seattle, Austin, etc. | |
| /zipcode | JSON:<br>{“value”: “**zipcode**”}<br><br>Zipcode: 02577, etc. | See JSON Output Object (v0.7) below |
| /fips | JSON:<br>{“value”: “**fips**”}<br><br>FIPS code: 01073000400, etc. | See JSON Output Object (v0.7) below |
| /coord | JSON:<br><br>{“latitude”: “**latval**”, “longitude”: “**longval**”}<br><br>Ex. For Boston:<br>Latitude: 42.361145<br>Longitude: -71.057083 | See JSON Output Object (v0.7) below |
| /county | JSON:<br>{“value”: “**fips**”}<br><br>5 digit county fips: 01073, 01083, ... | See JSON Output Object (v0.7) below |
| /hospitals | JSON:<br>{“state”: “**MA**”}<br>or<br>{“state”: “**MA**”. "county": "Suffolk"} | Hospitals, filtered by state, and optionally county |
| /hospitals/icu | JSON:<br>{“state”: “**MA**”} | ICU statistics by county |
| /batchstate | JSON:<br>{“value”: “**stateval**”}<br><br>stateval: MA, TX…<br>or a state name (Maine, Texas, etc)<br>or state FIPS (01, 02, 04…) | See JSON Output Object (v0.7) below |
| /batchcounty | JSON:<br>{“value”: “**stateval**”}<br><br>stateval: MA, TX…<br>or a state name (Maine, Texas, etc)<br>or state FIPS (01, 02, 04…) | See JSON Output Object (v0.7) below |
| /map/<state>/<city>/<zoom> | state: MA, TX…<br>or a state name (Maine, Texas, etc)<br>or state FIPS (01, 02, 04…)<br><br>city: Boston, Austin, etc.<br><br>zoom: between 1-18 (1 = the globe, 18 = close to street) | HTML Output that can be embedded in an iframe |

**JSON Output Object (Version 0.7)**

**Endpoint:**
/state

**POST Data:**
{“value”: “California”}
or
{“value”: “CA”}
or
{“value”: “06”}

**Output Example:**


```
[
  {
     "ARTHRITIS_CrudePrev": 0.181295,
     "BPHIGH_CrudePrev": 0.26661,
     "BPMED_CrudePrev": 0.664413,
     "CANCER_CrudePrev": 0.054201,
     "CHD_CrudePrev": 0.0,
     "COPD_CrudePrev": 0.0,
     "CSMOKING_CrudePrev": 0.138423,
     "DIABETES_CrudePrev": 0.097323,
     "HIGHCHOL_CrudePrev": 0.303718,
     "KIDNEY_CrudePrev": 0.029097,
     "OBESITY_CrudePrev": 0.249389,
     "STROKE_CrudePrev": 0.027094,
     "covid19_risk_score": 16.843503,
     "rank": 7,
     "robustness": "NA",
     "state": "CA",
     "version": "0.7"
  }
]
```


**JSON Batch Output Object (Version 0.7)**

**Endpoint:**
/batchstate

**POST Data:**
{“value”: “California”}
or
{“value”: “CA”}
or
{“value”: “06”}

**Output Example:**


```
[{"data": [
        {
           "ARTHRITIS_CrudePrev": 18.6,
           "ARTHRITIS_pred": 16.5,
           "BPHIGH_CrudePrev": 25.9,
           "BPHIGH_pred": 23.67,
           "BPMED_CrudePrev": 70.0,
           "BPMED_pred": 65.87,
           "CANCER_CrudePrev": 7.2,
           "CANCER_pred": 4.85,
            …
           "geoid": "06073017030",
```

```
        …
        "total_population": 23397
      },
      {
        "ARTHRITIS_CrudePrev": 21.8,
        "ARTHRITIS_pred": 18.76,
        …
      }
],
    "state": "CA",
    "version": "0.7"
}]
```


**Description:**

**covid19_risk_score**: XY's COVID-19 community risk score for the region (in this example, state)

**City_rank, state_rank, country_rank**: the rank of a particular entity (census tract, county, city, state, etc) relative to the next higher-up geographical grouping.

The **"_crudeprev"** fields are crude average weighted prevalences of conditions in a given area (state, city, etc) where: "*bphigh*" to high blood pressure, "*bpmed*" to prevalence of the population on blood pressure medication, "*chd*" to prevalence of heart disease, *csmoking* of the prevalence of the smoking, *highchol* to high cholesterol.

The **"_pred"** fields are the values predicted from XY's deep-learning pipeline.

The **"version"** field corresponds to the version of the API.

### 3. Using the Public API / Docker

To call an API endpoint, simple queries can be executed as follows (in Python 3):

```
import requests

# configure
requests.packages.urllib3.disable_warnings(requests.packages.urllib3.exceptions.InsecureRequestWarning)

# get data (change to https://localhost for docker)
res = requests.post('https://api.xyai-health.com/coord', json={"latitude": "42.361145", "longitude": "-71.057083"},
verify=False).json()

# print out a value
print(res[0]["COPD_CrudePrev"])
```

Data can also be easily accessed using cURL (one line):

```
curl -d '{"value": "MA"}' -H 'Content-Type: application/json' https://api.xyai-health.com/state
```
